# Critical dynamics predict cognitive performance and are disrupted by epileptic spikes, antiseizure medication and slow-wave activity

**DOI:** 10.1101/2024.08.19.24312223

**Authors:** Paul Manuel Müller, Gadi Miron, Martin Holtkamp, Christian Meisel

## Abstract

The brain criticality hypothesis postulates that brain dynamics are set at a phase transition where information processing is optimized. Long-range temporal correlations (TCs) characterizing the dissipation of information within a signal have been shown to be a hallmark of brain criticality. However, the experimental link between cognitive performance, criticality and thus TCs has remained elusive due to limitations in recording length, and spatial and temporal resolution. In this study, we investigate multi-day invasive EEG recordings of 104 persons with epilepsy together with an extensive cognitive test battery. We show that short TCs predict cognitive impairment. Further, we show that heterogeneous factors, including interictal epileptiform discharges (IEDs), antiseizure medications (ASMs) and intermittent periods with slow-wave activity (SWSs), all act directly to perturb critical dynamics and thus cognition. Our work suggests critical dynamics to be the setpoint to measure optimal network function, thereby providing a unifying framework for the heterogeneous mechanisms impacting cognition in conditions like epilepsy.

## Introduction

Cognitive function is an emergent property of cortical network structure and dynamics, and is often impaired by heterogeneous mechanisms in neuropsychiatric disorders [1–3]. In persons with epilepsy (PwE), cognitive impairment is a common comorbidity severely impacting daily functioning and quality of life [4]. PwE demonstrate a large heterogeneity in their cognitive function, with up to 30% of individuals having close to normal cognition [5]. The causes of cognitive dysfunction in epilepsy are multifactorial, stemming from both the underlying etiology of the disease, such as mesial temporal sclerosis, as well as dynamic factors including effects of antiseizure medications (ASMs), interictal and ictal activity, and disrupted sleep patterns [6,7]. Some of these factors, including interictal spikes, are known to also contribute to cognitive dysfunction in other diseases like Alzheimer’s [8–10]. Despite established associations of these factors, the neuronal mechanisms underlying cognitive function and heterogeneity in PwE remain largely not understood.

Central to intact cognitive function is the brain’s ability to process and integrate information across spatial and temporal domains [11–14]. Physics and information theory have provided a framework describing an optimal state of information processing. This critical state, poised at the phase transition between chaotic and ceasing neuronal activity, is characterized by an equilibrium between excitation and inhibition in the neuronal network [15–17]. When a network of neurons operates near a critical phase transition point, a range of information processing functions, including information transmission, integration, storage, dynamic range, and sensitivity to inputs, are optimized simultaneously [18–23].

The relevance of criticality in cortical networks is supported by experimental observations from animals and humans showing power-law scaling [24,25] and long-range temporal correlations (TCs) [21,26] which are hallmarks of critical network dynamics. Recent work has demonstrated that these signatures are predictably perturbed when networks are moved away from the critical point, either by external factors, such as ASMs [27,28], or internal influences, such as continued wake and sleep deprivation [27,29–32], providing further support for the relevance of critical dynamics in cortical networks.

While criticality provides a precise framework linking network structure to dynamics, its central claim, that critical dynamics predicts optimal network and thus cognitive function in humans, has not been proven yet. Using two unique, large-scale intracranial EEG datasets along with comprehensive cognitive testing, we here report that proximity to the critical point predicts cognitive impairment profiles in epilepsy. We show that heterogeneous factors, including interictal epileptiform discharges (IEDs), ASMs and intermittent periods with slow-wave activity (SWSs), all act directly to perturb critical dynamics and thus cognition. Our work suggests critical dynamics to be the setpoint to measure optimal network function, thereby providing a unifying framework for the heterogeneous mechanisms impacting cognition in conditions like epilepsy.

## Materials and Methods

### Participants

We conducted a retrospective analysis of persons with drug resistant focal epilepsy (PwE) that underwent presurgical evaluation by intracranial video-EEG monitoring. We validated our findings on two independent PwE cohorts. The first dataset was comprised of 81 PwE (35 females, average age 32.0 ± 10.7 years) assessed at the Berlin-Brandenburg Epilepsy-Center (BBEC / Dataset 1). All PwE underwent recording with subdural electrodes (SDE) and had an average of 57 ± 17 implanted electrodes per person. Forty-seven PwE had temporal lobe epilepsy (TLE) and 34 had extratemporal lobe epilepsy. All PwE underwent subsequent resection of the presumed epileptogenic zone, with 50 persons achieving good one-year post-surgical outcome (Engel score = 1). The second dataset included 23 PwE (12 female, average age 28.5 ± 13.2 years) from the epilepsy center at the University of Freiburg, available through the Epilepsiae database (ED / Dataset 2)[33]. PwE were implanted with subdural and depth electrodes (average electrode count 68 ± 27). Nineteen PwE had TLE and four had extratemporal lobe epilepsy, 20 underwent subsequent resection with 14 having good post-surgical outcome.

### Cognitive testing data

Cognitive testing was performed in all 81 BBEC subjects. Specifically, thirteen tests corresponding to four cognitive domains were performed during routine neuropsychological evaluation. Verbal learning and memory were evaluated using a word list learning test, the German version of the Rey Auditory Verbal Learning Test (“Verbaler Learn-und Merkfähigkeitstest”)[34]. Parameters assessed included immediate memory (VLMT1), reproduction of the last learning trial (VLMT5), total learning (sum of VLMT1-5) as well as delayed recall (VLMT7). Language was evaluated using semantic and phonetic word fluency tests with and without category switching (“Regensburger Wortflüssigkeitstest”)[35]. Working/short term memory was assessed by two block tapping tests and Mottier tests; and attention was assessed using the computerized Test battery for Attentional Performance (TAP; “Testbatterie für Aufmerksamkeitsprüfung”) with the subtests Alertness and Go/No-Go [36–38]. All test scores were normalized regarding age and sex according to established norms provided in the test manuals and converted into percentile ranks. If single test scores were missing those were iteratively imputed using *scikit-learn*’s iterative imputer [39]. Cognitive domain impairment was defined as two parameters within a domain having a score of at least one standard deviation (SD) below the mean of the norm population.

### Quantification of antiseizure medications

In the BBEC dataset (dataset 1), charts were manually reviewed, and all antiseizure medications (ASMs) given during video-EEG monitoring were extracted. In the ED (dataset 2), daily ASM dosing information was provided as part of the documentation. For each day, a daily dose was defined by normalizing medications by their individual defined daily doses. For each patient that underwent ASM tapering as part of the clinical video-EEG monitoring procedure, days with the highest and lowest dose were identified according to the summed prescribed drug dosages normalized by their individual defined daily dosages across drugs (high and low ASM days). Days with rescue medication (midazolam, diazepam, or lorazepam) were excluded due to their strong neurophysiological effects. Subjects that did not undergo medication tapering were excluded from analyses related to ASM effect.

### Preprocessing of intracranial electroencephalography data

In the BBEC dataset, intracranial electroencephalography (iEEG) data was available for the first 5 minutes of each hour during the multi-day monitoring period, whereas in the ED, iEEG data was available continuously. Analyses were performed on the first and last day of full recording, and on the high and low ASM dosage days. Pre-processing of iEEG was applied in the following order: first, notch-filtering was applied to remove power line noise at 50 and 100 Hz; second, signals were down-sampled to a common frequency of 256 Hz (original sampling frequencies varied between 256 – 2048 Hz) by using an anti-aliasing filter and then decimating the signal; third, bandpass-filtering from 0.1 to 128 Hz was done to remove slow drifts and high frequency artifacts; fourth, channels with constant signal or abnormal frequency peaks (> 6 times interquartile range across the full recording) were removed; lastly, visual inspection was performed to verify the preprocessing and remove additional clearly artifactual channels, if needed. Seizure segments together with at least 10-minute pre- and post-ictal periods were excluded from the analysis; EEG seizure on- and offset had been marked by clinical experts.

### Temporal correlations calculation

Temporal correlations (TCs) have been shown to characterize information processing, and their maximisation is a hallmark for criticality [29,40,41]. Here, TCs are calculated as previously described [27,29]. In detail, power fluctuations in the high *γ*-band are evaluated as they most closely capture local spike rate variations [42–46]. For this purpose, the median spectral power from 56 to 96 Hz is calculated every 125 ms using Welch’s method (Hann window). Next, powers are normalized through applying the logarithm with base 10. Then, autocorrelation functions of the high *γ*-power time series are calculated in consecutive 120-second windows (90 seconds overlap) for each iEEG channel. The decay of the autocorrelation function (ACF) is quantified by TCs, which are defined as the first time the autocorrelation function drops below half the difference between the first lag value and a baseline, i.e., the median of the ACF between 40 and 60 seconds. The ACF value at lag zero is excluded as it is one by definition and thus independent of inherent noise and data quality levels, and its inclusion may obscure the true underlying ACF decay rate. As a result, the minimal value for TCs is 125 ms. To ensure robustness of the TCs, ACFs are first aggregated (median) over multiple 120-second windows. In case of assessing global states, e.g., comparing low and high ASM days, we subsampled to the number of time points in the minority group. In the case of channel specific states, e.g., assessing TCs as a function of IED load, we subsampled to 50 points as a trade-off between robust TCs calculation, maximization of channel inclusion and comparability between channels. All subsampling procedures were repeated 100 times by random shuffling, and average values over the repeats are reported. As a baseline surrogate TCs were calculated from time shuffled high-*γ* power time series. This procedure keeps the power distribution constant but destroys the temporal relation of the data points.

### Detection of slow waves

Apart from slow-wave sleep, spatially and temporally localized slow waves also emerge as local sleep-like activity within the awake brain where they accompany behavioral markers of lapses and precede reports of mind wandering and mind blanking [47]. We therefore classified periods with slow-wave activity (slow-wave sleep, SWS) across the full wake-sleep continuum. SWS was scored on 30-second windows using a validated algorithm [48]. First, a vigilance index is calculated as the fraction of the powers

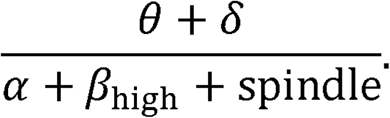

Second, SWS was defined as one standard deviation over the mean for each day separately [49]. In order, to align these 30-second segments with the TC analysis based on 120-second segments, we chose to define a 120-second window to be marked as ‘SWS’ if at least one of the four 30-second segments in it was identified as SWS. Otherwise, the 120-second segment was labelled ‘nonSWS’.

### Detection of interictal epileptiform discharges

Interictal epileptiform discharges (IEDs) were identified using an automated deep-learning method that has previously been described and validated [50]. First, a template matching filter was applied to identify possible IEDs activity within each channel separately. Next, the raw EEG signal was transformed into spectrograms to reduce the dimensionality of EEG features. This signal was then subjected to a pre-trained deep neural network for intracranial IED detection. To analyze the effect of IEDs on TCs, we compared TCs between the following IED per minute count bins: no IEDs, (0-1], (1-5], and (5-30] IEDs per minute.

### Spectral power calculation

Spectral powers were used as control measures when evaluating the relationship between TCs and cognitive impairment. We calculated the median power for every 120 second segment in the following bins using Welch’s method (Hann window): *δ* (0.5-4 Hz), *θ* (4-8 Hz), *α* (8-12 Hz), *β* (12-30 Hz), *γ* (30-45 Hz) and high-*γ* (55-95 Hz).

### Statistical analysis

For analyses comparing states of the same patients (ASM, SWS, and between IED bins) we used paired Wilcoxon. The influence over all IED bins on TCs was evaluated using a linear mixed effects model: dependent variable: TC; independent variables: ordered IEDs, SOZ or nSOZ, interaction of term SOZ and IEDs; random effect: patient ID.

In order to compare significance across patients with respect to cognition, we applied the Brunner-Munzel tests as distributions of TCs are non-normal and heteroscedastic. We corrected for multiple comparisons using the Benjamini-Hochberg method at a corrected *α* < 0.05. Additionally, we report effect size through the non-parametric relative effect. Lastly, the distribution of p-values is compared with a Kolmogorov-Smirnov test to a uniform distribution, as one would expect under the null hypothesis.

### Neural network modelling

To guide our investigation of TCs, we studied a parsimonious neural network model, whose distance to criticality can be tuned by one parameter, and which has built-in mechanisms modelling ASM effects, SWS, and IEDs, similar to the model in [27–29]. The model consists of *N* = 1024 neurons on a 2-dimensional equidistant grid with periodic boundary conditions. Neurons are all-to-all connected with random uniform strengths scaled by a distance-dependent Gaussian profile

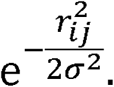

The Euclidian distance between neuron *i* and *j* is *r*_*ij*_ and *σ* is the scaling width of the profile which was set to *σ* = 4 as in [51,52]. We omitted self-connections and randomly set 20% of neurons to be inhibitory and 80% to be excitatory, altogether resulting in the weight matrix 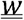. The general strength of the weights is scaled based on the largest absolute eigenvalue *λ* of this weight matrix, where *λ* = 1 corresponds approximately to the transition point between ceasing and runaway activity, i.e., between sub- and supercritical dynamics (*λ* < 1 and *λ* > 1, respectively) [17,53].

Each neuron can at time *t* either be active *s*_*i*_ (*t*) = 1 or inactive *s*_*i*_ (*t*) = 0. At the next time step *t* + 1 a neuron will become active based on its inputs with probability

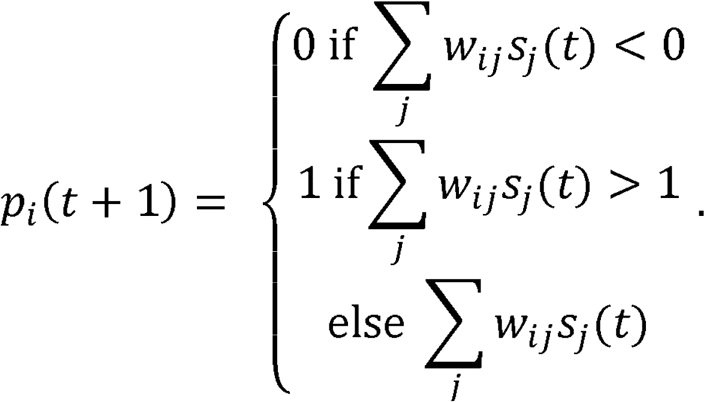

Additionally, one neuron is set to be active at each of the max_*t*_ = 5000 time steps as background activity. For the calculation of TCs, the first 500 timesteps are discarded to account for the initial transient.

Following previous work, the influence of ASMs is modelled by reducing the outgoing excitatory connection strengths by a factor *f*_exc_, motivated by the mechanism of ion-channel blockers [28]. Deep slow-waves sleep (SWS) is represented by introducing off-periods at random times where all neurons remain quiescent with probability *p*_Off_ [29]. To model IEDs, we introduced local perturbations by setting a random neuron and its local neighbours (20% of all neurons) to be active with probability *p*_IED_. Model simulations are repeated 1000 times. TCs were extracted from the autocorrelation function of the average neuron firing.

### Ethics statement

This study was approved by the Institutional Review Board of Charité – Universitätsmedizin Berlin. Due to the retrospective nature of the study, informed consent of patients was waived. The use of patient data from the Epilepsiae database was approved by the Institutional Review Board of the University of Freiburg and written informed consent that the clinical data might be used and published for research purposes was given by all patients [61].

### Data and code availability

Code for the model will be made available at https://gitlab.com/computational-neurologie/stc_model_2024 at the time of publication. Code for the analysis will be made available under https://gitlab.com/computational-neurologie/sde_n_cog for the BBEC dataset and https://gitlab.com/computational-neurologie/ieegCD for the ED. The ED dataset is publicly available within the Epilepsiae database [61].

## Results

We first demonstrate that heterogeneous mechanisms known to act on cognition, including antiseizure medications (ASMs), interictal epileptiform discharges (IEDs) and intermittent periods with slow-wave activity (SWSs), all act to perturb the optimal critical network state as a common target. Second, we show that the proximity to this critical network state is the strongest predictor of cognitive performance, thus suggesting it to be the setpoint to monitor cognition.

### Disruption of optimal critical network state by heterogeneous mechanisms in a network model

We first analyze a parsimonious neuron network model based on a branching process [27] to review how optimal collective network dynamics are shaped by network interactions and perturbed by different mechanisms known to act on cognition (ASMs, IEDs, SWSs). Analyzing the model over varying connection strengths revealed the well-known phase transition when the largest absolute eigenvalue of the weight matrix *λ* approached one (Figure 1 A). For *λ* ≪ 1 the network was in its subcritical regime, activity of the neurons ceased, and TCs were shortened (Figure 1 D). For *λ* ≫ 1 the system was in its supercritical regime, the activity exploded, TCs remained short. At *λ* ≈ 1 the system was in its critical regime, and self-similar activity emerged. Here, TCs exhibited maximal duration, showing a state where information could stay in the system for the longest time (Figure 1 D.) We next investigated the effects of SWSs, IEDs and ASMs on critical dynamics.

**Figure 1:**
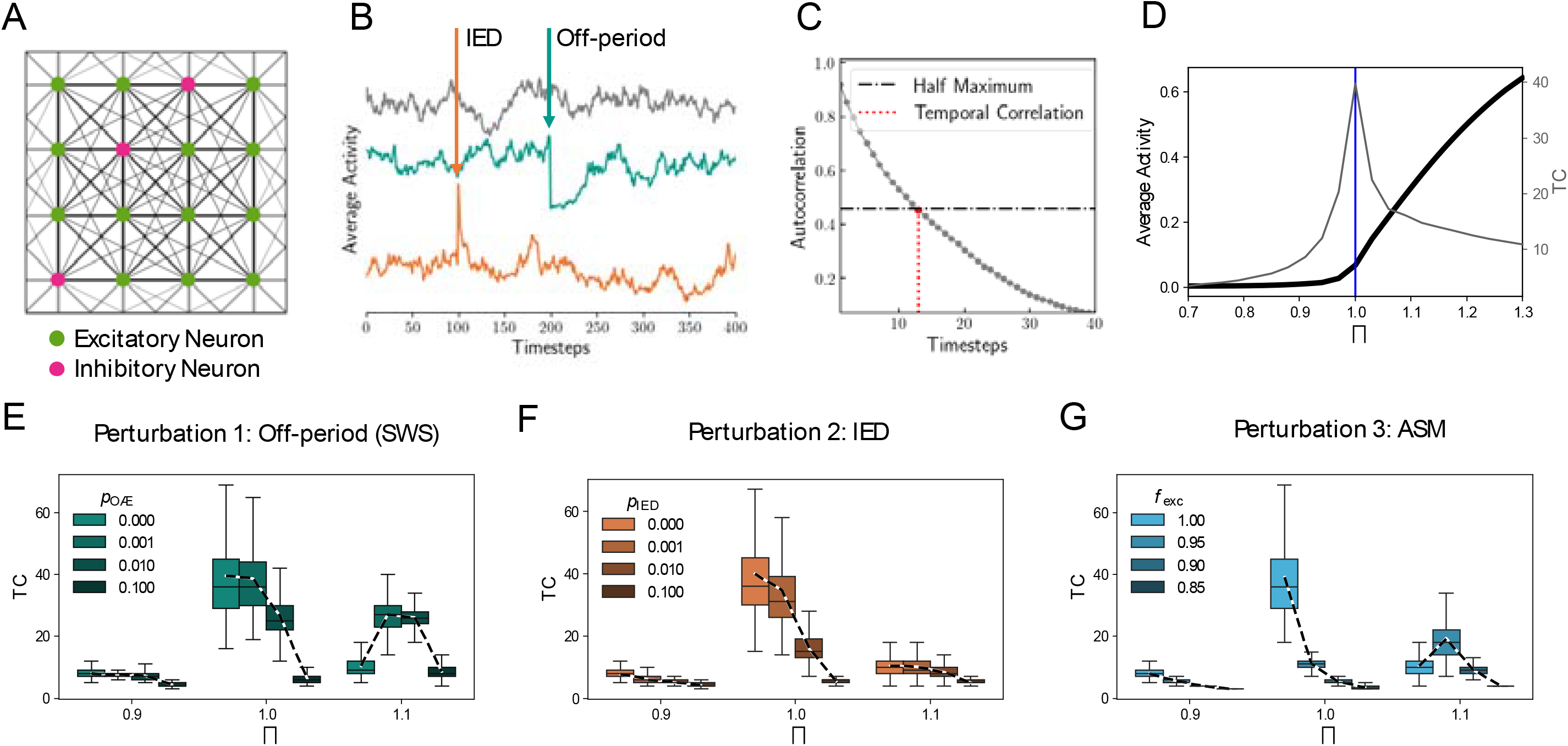
Neuronal network simulations indicate maximal TCs around the critical point, and their decline under SWSs, IEDs and ASMs. A, Networks of size *N* = 1024 (20% inhibitory 80% excitatory neurons) are coupled with distance dependence on a grid with period boundary conditions. B, Unperturbed model run at criticality (grey), with an off-period (at timestep 200, teal), with an IED (at timestep 100, orange). C, Definition of TC as the half width at half maximum of the autocorrelation function. D, Phase transition of the order parameter (average activity, black) at which TCs (grey) peak. E, TCs as a function of SWS in the subcritical (*λ* = 0.9), critical (*λ* = 1) and supercritical (*λ* = 1.1) regimes. F, TCs as a function of IEDs in all dynamical regimes. G, TCs as a function of ASMs.

First, we modeled SWS by introducing the possibility that neurons could synchronously go offline with probability *p*_Off_ as in [29] (Figure 1 B). For the three dynamical regimes of the model, i.e., subcritical, critical, and supercritical, TCs declined under large p_Off_ (Figure 1 E). Only in the supercritical regime, an increase of TCs for small *p*_Off_ can be observed as effectively transients towards saturated dynamics are introduced. We observed the strongest decline in TCs if the system was initially close to the critical regime, *λ* =1.

Second, we introduced IEDs by the activation of local populations of neurons with probability P_IED_ (Figure 1 B). For all model regimes, increasing *p*_IED_ led to shortening of TCs, with the strongest effect seen again in the critical regime (Figure 1 F).

Third, we simulated the effect of ASMs by reducing the connection strength of excitatory neurons by factor *f*_exc_ (Figure 1 G). In the subcritical and critical regime, we found a reduction of TCs with decreasing *f*_exc_. In the supercritical regime (*λ* = 1.1), slight reductions of excitability lead to longer TCs, whereas larger decreases of *f*_exc_ also lead to lower TCs.

Collectively, model results provided testable hypotheses. In line with previous work, they indicated that optimal information processing should be found around a critical point where TCs are maximized. Conversely, SWSs, IEDs and ASMs all shifted dynamics away from criticality, thereby reducing TCs as a potential signature of cognitive impairment.

### Disruption of optimal critical network state by heterogeneous mechanisms in human iEEG

To test these hypotheses, we investigated TCs from iEEG data of 104 PwE as a function of SWSs, IEDs and ASM load (Figure 2 A, B).

**Figure 2:**
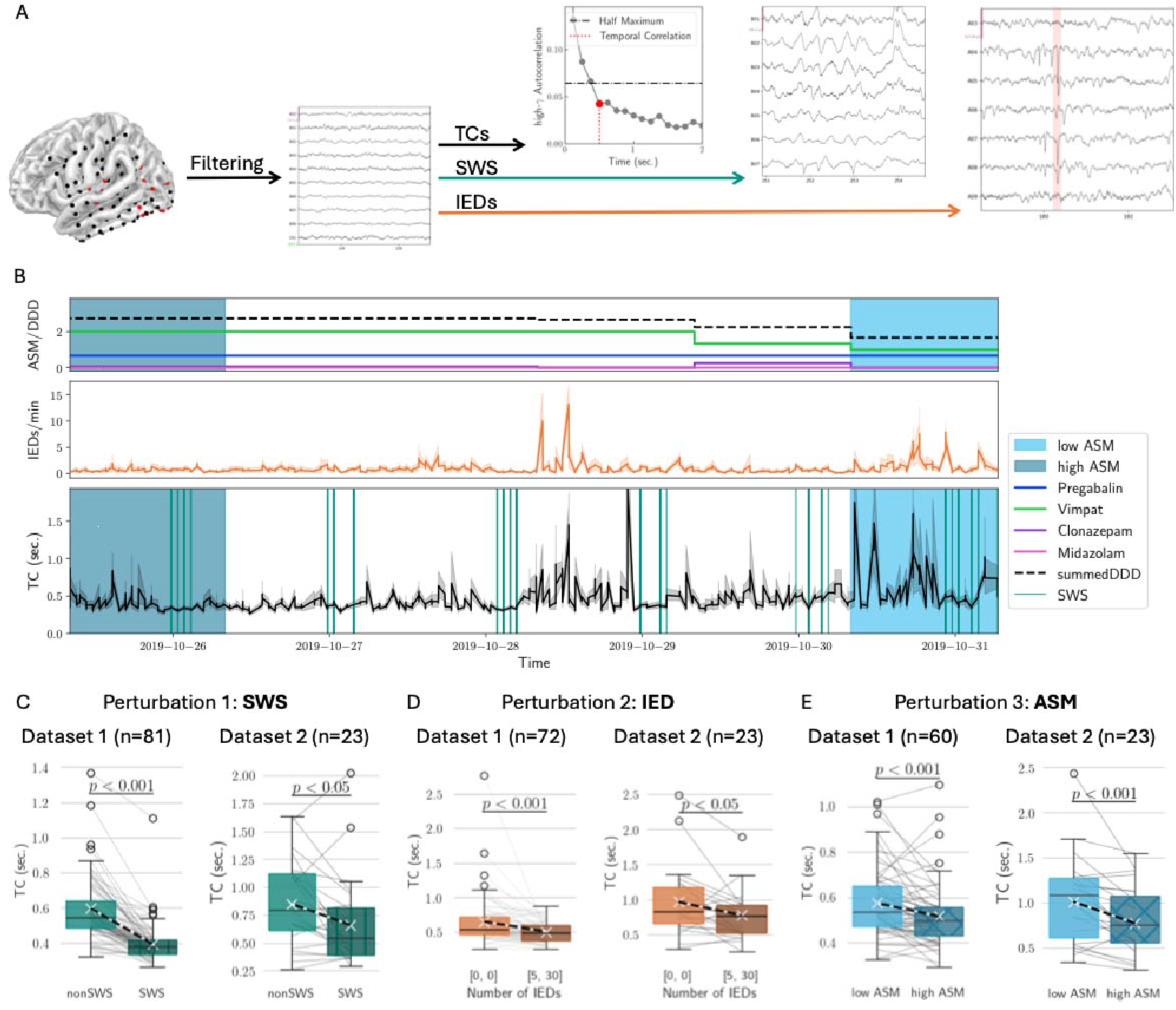
Disruption of TCs and network optimality through SWS, IEDs and ASMs. A, iEEG data from persons with epilepsy followed by extraction of TCs and classification of segments with SWSs and IEDs. B, For each patient, ASM dosage was recorded and aligned with IED rate, SWS segments and TCs. C, TCs as a function of SWS in the two datasets. D, TCs as a function of IEDs. E, TCs as a function of ASMs.

First, SWS epochs were identified through a validated algorithm [49]. In the BBEC dataset (dataset 1) (18 ± 7)% (mean ± standard deviation) of the segments per patient were classified as SWS (19 ± 6)%and (in the ED dataset. In both independent datasets, we found TCs during SWS significantly shortened compared to nonSWS segments (BBEC: TC_non SWS_ = (0.6 ± 0.2) sec. vs TC_SWS_ = (0.4 ± 0.1) sec., *p* < 0.001; ED: TC_non SWS_ = (0.9 ± 0.4) sec. vs TC_SWS_ = (0.7 ± 0.4) sec., *p* < 0.05; Figure 2 C).

Second, we investigated the effect of IEDs on TCs. Using a validated algorithm [50], we identified an average of (2.6 ± 2.4) IEDs/(channel·min) across subjects in the BBEC dataset and (3.1 ± 1.7) IEDs/(channel·min) in the ED dataset. In both datasets, longer TCs were observed in the absence of IEDs (BBEC: TC_no IED_ = (0.6 ± 0.4) sec. vs TC_[5.30] IEDs_ = (0.5 ± 0.2) sec., *p* < 0.001; ED: TC_no IED_ = (0.9 ± 0.5) sec. vs TC_[5,30] IEDs_ = (0.7 ± 0.3) sec., *p* < 0.05; Figure 2 D).

Third, we evaluated the effect of ASMs on TCs by comparing high and low ASM dosage days for the 60 BBEC and 23 ED subjects that underwent ASM tapering during monitoring. BBEC patients had an average ASM load reduction of (50 ± 30)% from high to low ASM days, ED patients of (70 ± 30)%. In both datasets, TCs were shortened under increased ASM loads (BBEC: TC_low ASM_ = (0.6 ± 0.2) sec. vs TC_high ASM_ (0.5 ± 0.1) sec., *p* < 0.001; ; ED: TC_low ASM_ = (1.0 ± 0.5) sec. vs TC_high ASM_ (0.8 ± 0.4) sec., *p* < 0.001; Figure 2 E). None of these effects for SWSs, IEDs and ASMs were observable for the surrogate TCs.

### Optimal network state disruption inside and outside the seizure onset zone

The seizure onset zone (SOZ) refers to the brain region(s), where seizures originate. We next examined the network state and the effects of these three perturbations separately in the SOZ and non-seizure onset zone (nSOZ) electrodes (Figure 3). We found TCs similarly shortened in the nSOZ and the SOZ as a function of SWSs (Figure 3 A, D) and ASMs (Figure 3 C, F) demonstrating the widespread network effects incurred by these perturbations.

**Figure 3:**
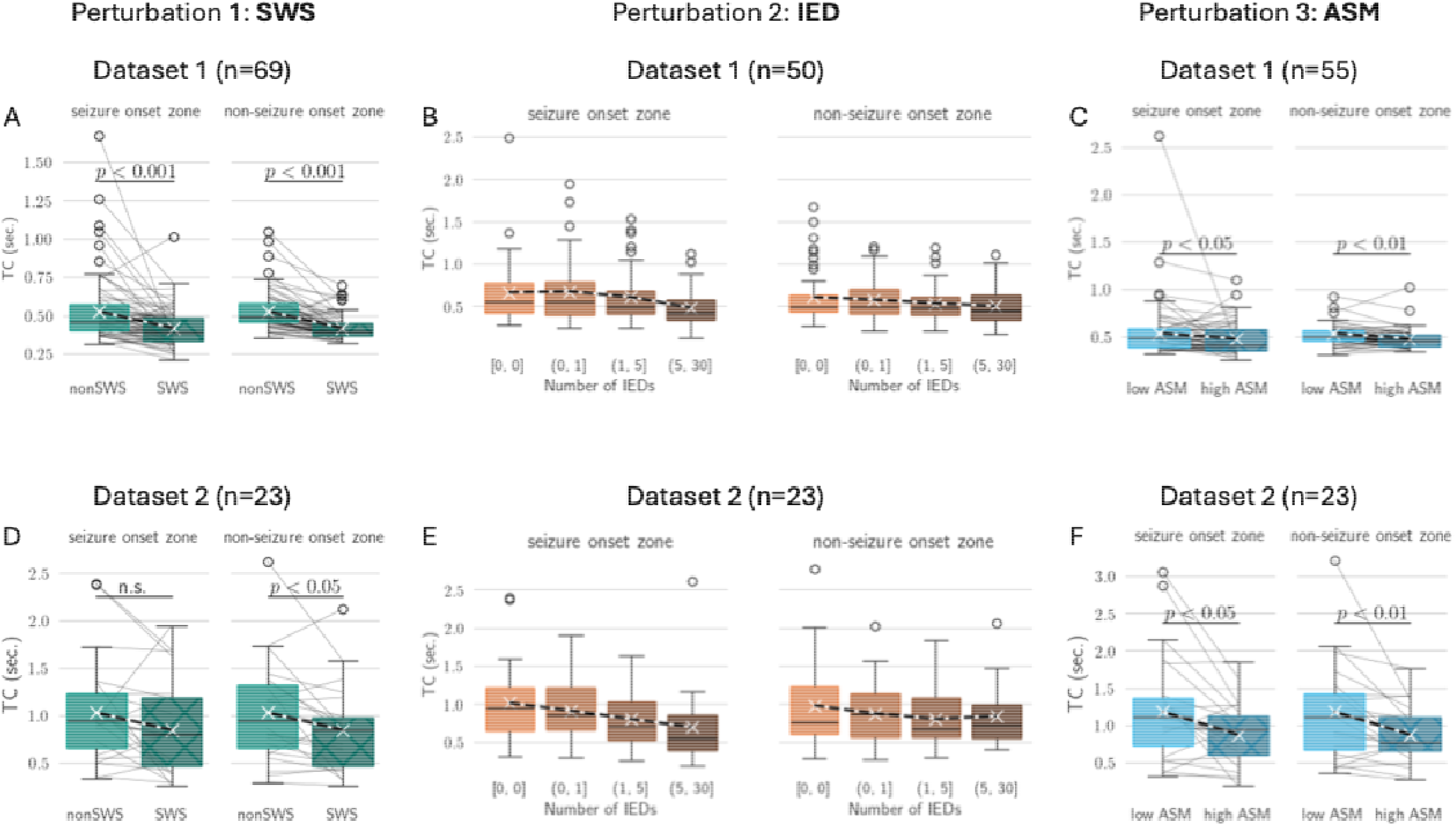
TCs decline under perturbations in the seizure onset zone (SOZ) and non-seizure onset zone (nSOZ). Dataset 1: A, SWSs disrupt TCs both in the SOZ and the nSOZ. B, With increasing number of IEDs, TCs progressively decline in both SOZ and nSOZ. C, TCs decline with increased ASM load in SOZ and nSOZ. Dataset 2: D, E, F.

Similarly, in both datasets, we observed that TCs were progressively shortened under increasing numbers of IEDs in the SOZ and nSOZ (Figure 3 B, E). For this analysis, to allow comparability across patients, we included only PwE who had at least 50 segments in each IED bin for at least one channel, excluding SWS segments. For the BBEC dataset (comprised of 50 eligible PwE), we found negative linear and quadratic IED coefficients (linear: -0.13 [-0.20, -0.07] [Wald confidence intervals], t=-3.9; quadratic: -0.069 [-0.0136, -0.02], t=-2.0). The former shows that with increasing IEDs TCs were progressively shorter and the latter indicating that this decline was decelerating for higher IED counts. Additionally, the model revealed that nSOZ areas, compared to SOZ areas, had smaller baseline TCs (−0.052 [-0.997, - 0.02], t=-2.1). This effect was less marked in the ED dataset, where all 23 patients were eligible. Here, only the linear decrease in TCs with increasing IED bin was robust (−0.24 [-0.35, -0.13], t=-4.1), with no robust effects discerning the SOZ from the nSOZ. The prediction of the models is illustrated in Supplementary Figure 1. Together these observations thus indicate a more rapid decline of TCs as a function of IEDs in the SOZ and overall higher baseline TCs in the SOZ.

### Distance to optimal critical dynamics predicts cognitive impairments

In model and data, IEDs, ASMs and SWSs all acted to perturb critical dynamics. This suggests critical dynamics to be the common setpoint for optimal network function. We thus finally assessed whether distance to criticality predicted cognitive performance across all heterogenous factors. All PwE undergoing presurgical evaluation at the BBEC underwent cognitive assessment in the language, verbal learning and memory, working memory, and attention domains. Subjects were classified based on their performance in each domain as either conserved or impaired cognition. We investigated the relationship between TCs and cognitive performance, adjusting for the varying number of recording days by analyzing the first, last, and lowest ASM load days. To account for the spatial variability of TCs across the cortex, sub-analyses were performed by hemispheres and lobes. To mitigate false positives, signal-related artifacts, and general EEG signal changes, we parallelly conducted analyses for surrogate TCs, standard spectral band powers, existence of lesion, IED counts, SWS count per day, and ASM levels (Figure 4 A).

**Figure 4:**
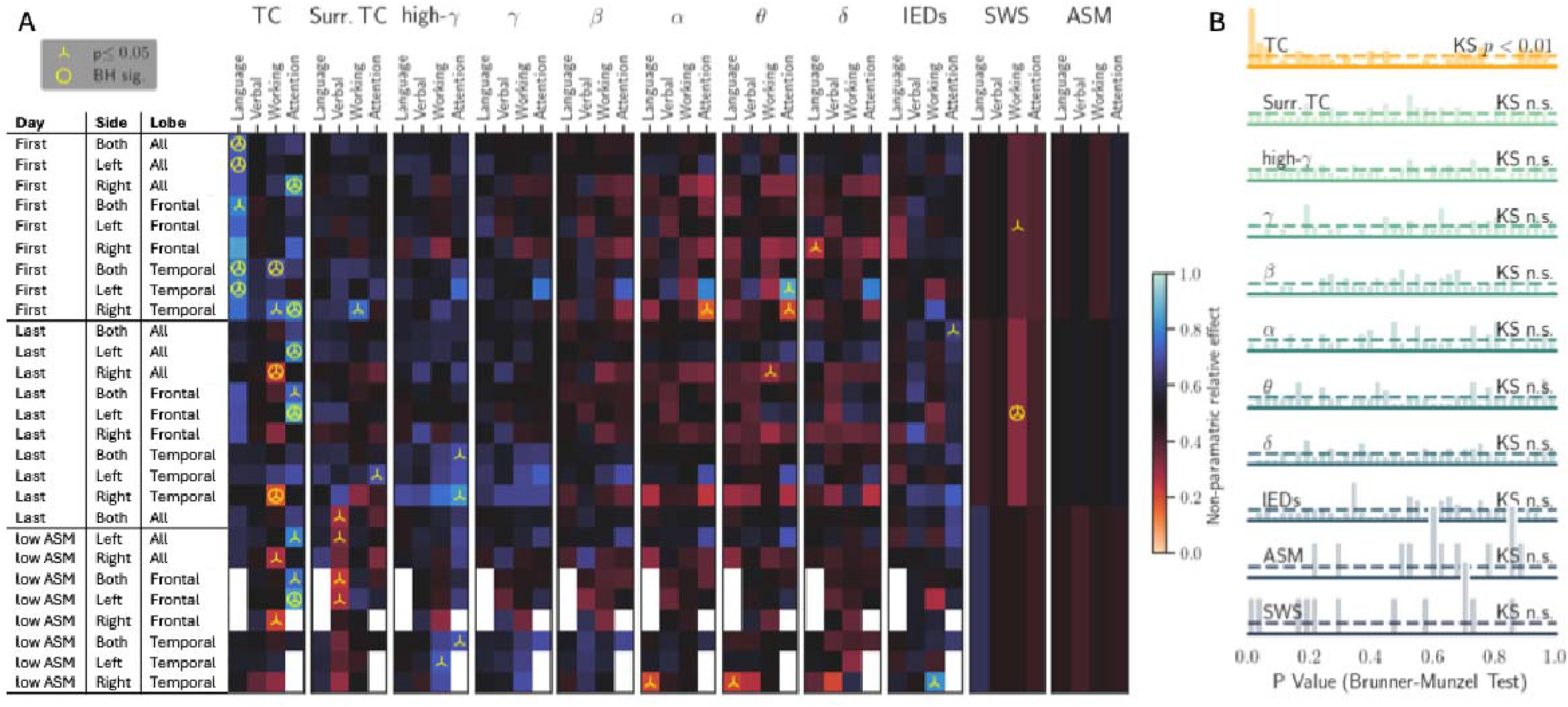
Cognitive impairment is predicted robustly only by TCs. A, EEG and clinical features were compared to performance on cognitive tests, each box denotes one comparison. On the left, the table notes the temporal and spatial characteristics of sampling. We assessed EEG features during the first, last and low ASM days, across temporal and frontal lobes, and both hemispheres. Each heatmap contains four columns in order to relate EEG features to performance in specific cognitive domains: language, verbal learning and memory, working memory, and attention. A cognitive domain was considered impaired if two or more tests within the domain were 1 standard deviation under the healthy norm. Each heatmap column group refers to a single EEG feature examined: temporal correlations (TCs) and surrogate TCs, power spectra in the *δ* (0.5-4 Hz), *θ* (4-8 Hz), *α* (8-12 Hz), *β* (12-30 Hz), *γ* (30-45 Hz) and high-*γ* (55-95 Hz) ranges, and IEDs. In addition, SWSs and ASM load were investigated. The color within each box denotes the effect size, statistical significance is noted by a triangle at *p* < 0.05 and by a circle after Benjamini-Hochberg correction. White boxes had insufficient sample size for analysis. B, P-values of all comparisons.

We identified a significant relationship between TCs and cognitive impairment, with 19 out of 102 comparisons showing significance (*p* < 0.05, Brunner-Munzel test; Figure 4 B), of which 12 retained significance after adjusting for multiple comparisons (Benjamini-Hochberg method with *α* = 0.05). TC shortening was the only measure notably correlated with impairments in attention and language; no other EEG features were significant after multiple comparison correction (Figure 4 A). Importantly, neither IEDs, ASM load or SWSs (with the exception of one value, as a non-localized measure only 12 comparisons could be performed) exhibited significant predictive value for cognition (Figure 4 A). Additionally, the existence of lesions in this PwE cohort as detected by MRI did not correlate with any cognitive domain (*p* > 0.05). In line with these observations, a broader analysis of the p-value distributions for all features revealed that only TC’s p-value distribution significantly deviated from a uniform distribution expected under the null hypothesis (*p* < 0.01, one-sided Kolmogorov-Smirnov test; Figure 4 B).

## Discussion

Despite receiving considerable attention, the crucial link to cognitive function predicted by the brain criticality hypothesis has so far remained unproven [54–57]. We address this gap in research by demonstrating that TCs, a hallmark of critical dynamics, predict cognitive performance. We show that heterogeneous factors known to impair cognition, including IEDs, ASMs and intermittent periods with slow-wave activity (SWSs), all act directly to perturb critical dynamics and thus cognition. Our work suggests critical dynamics to be the setpoint to determine optimal network function, thereby providing a unifying framework for the heterogeneous mechanisms impacting cognition in conditions like epilepsy.

Cortical networks have long been hypothesized to operate in the vicinity of a critical state where they can benefit from optimized function by balancing efficient information processing, memory and flexibility. Sleep has been shown to play a crucial role in retuning cortical networks to this optimal critical state [29,30,31]. TCs in neuronal dynamics measure information dissipation and serve as an indicator of this optimal state [58,59,60,61,62]. Thus, disrupted TCs indicate a deviation from the optimal critical regime. Despite theoretical arguments connecting information processing with criticality, experimental evidence has been lacking. This is in part due to limitations related to EEG recording settings, including short EEG recording durations inadequate to capture the statistical properties of critical dynamics, coarse spatial brain coverage of electrodes, artifacts and signal filtering induced by scalp EEG, and comparisons made to single cognitive test measures which may not accurately reflect overall cognitive performance. In particular, it is crucial to capture high-*γ* power fluctuations which closely capture local spike rate variations [42–46] because, consequently, only TCs in high-*γ* power change predictably and robustly under ASM titrations or during SWS in contrast to TCs obtained from lower frequencies, e.g., *α*-band TCs (see Suppl. Fig.2 in [27]). These high-*γ* power frequencies are typically filtered out by the scalp in scalp EEG and are even less resolved in slowly fluctuating signals like the BOLD signal. Hence, markers of criticality could not been connected robustly to cognitive performance measures in fMRI [58] or scalp EEG research [59].

We overcome this limitation here by using iEEG, analyzing high-*γ* TCs from 104 PwE in long-duration recordings of one week on average with high spatial resolution and compared them to outcomes of an extensive, standardized and clinically validated cognitive test battery of 13 tests spanning four different cognitive domains. Our specific hypothesis, based on criticality theory and results from our computational model, was that disrupted TCs are a signature of cognitive impairment. Our results indicate that TCs are robustly shorter for patients with cognitive impairments. Notably, the domains that were found to be related to TC disruption were language and attention. Although we demonstrate that cognitive profiles relate to TC duration, the effects on cognition are well known to be multifactorial and highly dynamic. While cognitive phenotyping was performed according to best-practice clinical standards, it still reflected an assessment that was done at a single point of time. This is a common limitation of experimental studies comparing cognition to brain function in general [56].

Our results shed light on the underlying cognitive impairments in conditions like epilepsy which have remained largely elusive. Specifically, we examined mechanisms known to affect cognition: effects related to intermittent periods of local or global SWS, increased IED activity and increased ASM load. We find that perturbations to the system introduce a disruption of TC. SWS was simulated in our model by introducing off periods, where neuronal activity is markedly reduced. This approach was based on experimental work showing off-periods in rats during sleep [60] and extended wakefulness [29,61]. Our model predicted that during SWS there is a shift of the system toward subcritical dynamics, with shortening of TCs. These predictions were validated here on two independent datasets. Our findings align with theories that during sleep and sleep-like periods the brain loses its ability to effectively integrate information across cortical areas and time [62]. Further, our results relate to prior studies that have shown that extended wakefulness shows impaired cognition possibly caused by intermittent local sleep activity [29,30,63]. In PwE, this is an especially pertinent consideration as epilepsy disturbs sleep through multiple mechanisms, influencing sleep structure, architecture, continuity and oscillations [64]. Subsequently, PwE are more prone to the effects of sleep disturbances contributing to cognitive impairments.

IEDs, in contrast, were simulated by probabilistic activations of local neuronal population. Our computational model predicted IEDs would disrupt TCs, and this was confirmed in both experimental datasets. Notably, we also measured a dose-dependent effect, with increased IED rates resulting in shorter TCs. This suggests that increased disease activity is directly related to a shift away from the critical state. This finding may provide a mechanistical perspective to the previously reported ‘transient cognitive impairment’ phenomenon, which describes impaired cognitive performance due to concurrent epileptic activity during cognitive testing [65,66]. It may also potentially shed some light on the neuronal mechanisms underlying impaired cognition associated with IED activity reported in neurodegenerative disorders including Alzheimer’s disease [67]. Although we demonstrate a drift away from criticality with increased IEDs, we cannot draw direct conclusions if the deviation is towards sub- or supercritical dynamics. One prior study suggested that IEDs themselves are a display of supercritical dynamics [68], however additional work is needed to resolve this open question.

The effect of ASMs was simulated by reducing the strength of excitatory connections between neurons, similarly to the mechanism of ion channel blockers. We demonstrated that both in model simulations and experimental data, increased ASM dose shortened TCs, extending initial findings from prior works [27,28]. The relationship between ASMs and cognition has been well-documented [69]. ASM treatment is recognized to have a major dose-dependent detrimental effect on cognition in PwE, limiting adherence towards treatment and negatively affecting quality of life[69]. While there are over 20 different ASMs that function through distinct mechanisms, we aimed to examine a general effect that can be seen across treatments. One hypothesis is that the reduction of neuronal excitability through ASMs [70,71] can lead to decreased effective connectivity between the neurons and thus a transition towards subcritical dynamics and consequentially impaired cognition [72].

Findings of the effects of perturbations were consistent across both seizure and non-seizure onset tissue, suggesting our findings may not only be an epilepsy-specific effect. Only in the BBEC dataset, we found a stronger decline of TCs with increasing IEDs in the SOZ compared to the nSOZ. Further, correcting for IEDs in the SOZ revealed slightly larger TCs than in the nSOZ. This finding may support the hypothesis that the SOZ is closer to criticality and thus is more prone to drifting into a supercritical state, a phenomenon which is also linked to seizure initiation [73,74]. To note, in the ED dataset we could not establish this finding, possibly due to a smaller sample size, similarly to a prior study [75].

Our study has several limitations. First, we had continuous recordings only for the subjects from the ED. In the BBEC dataset, we were limited to sampling 5 minutes each hour. However, PwE were recorded for relatively long durations, and compared to similar datasets, we had a very large sample size of over 100 patients. Second, due to the large sample size it was not feasible to manually annotate IED segments. We used a well-validated algorithm for this purpose, however naturally there could be misclassified segments. Third, it was not possible to perform sleep scoring according to polysomnographic standards as PwE did not have EMG or scalp EEG recordings, however SWS segments were derived using a validated algorithm designed for iEEG recordings. Lastly, cognitive testing was done as part of the routine clinical workup, and thus was not concomitant with iEEG. This can lead to bias due to the dynamic nature of cognitive impairment in epilepsy, as well as differences in ASM dosage during cognitive testing. However, we examined a clinically robust battery of cognitive tests in order to establish the cognitive profiles of all subjects. We employed multiple measures to verify the robustness of our results. First, in addition to TCs, we analyzed frequency band powers and IED rates. These were all shown to be non-significant when compared to cognitive outcomes. Non-significance of the high-*γ* power band is especially a strong indicator that artifactual or other general signal properties do not explain our findings, as TCs were derived from gamma power band signal. The fact that the IED rates were not significantly related to cognitive results further indicates that we did not merely measure an effect related to disease severity or activity. Second, we compare our results to chance by statistically demonstrating that our findings are different from the expected uniform distribution if the null hypothesis is true. Third, we show our results remain robust with multiple testing corrections, despite the high number of tests performed. Fourth, to control for different monitoring durations among patients, we perform multiple analyses accounting for the day of recording. Notably, we found a statistically stronger relationship between TC to cognitive impairment in the first day of recording. This may be due to the other days of comparison introducing large amounts of variability in the data as PwE having different monitoring durations, variable drug tapering schedules, and other accumulating biases. Nevertheless, significant findings were also demonstrated on the last recording days and lowest ASM dose days. Finally, we also compared cognitive outcomes to SWS and ASM dosage levels, excluding these external influences as systemic biases to our findings.

Collectively, our results from model simulations and intracranial EEG recordings show that TCs decline under perturbative mechanisms – SWS, IEDs, and ASM – and that shorter TCs predict cognitive impairment in PwE. More generally, our results demonstrate how dynamical factors may distinctly influence the brain’s distance from a critical state and thereby cognition, as illustrated in Figure 5. Our work suggests critical dynamics to be the setpoint to measure optimal network function, thereby providing a unifying framework for the heterogeneous mechanisms impacting cognition in conditions like epilepsy.

**Figure 5:**
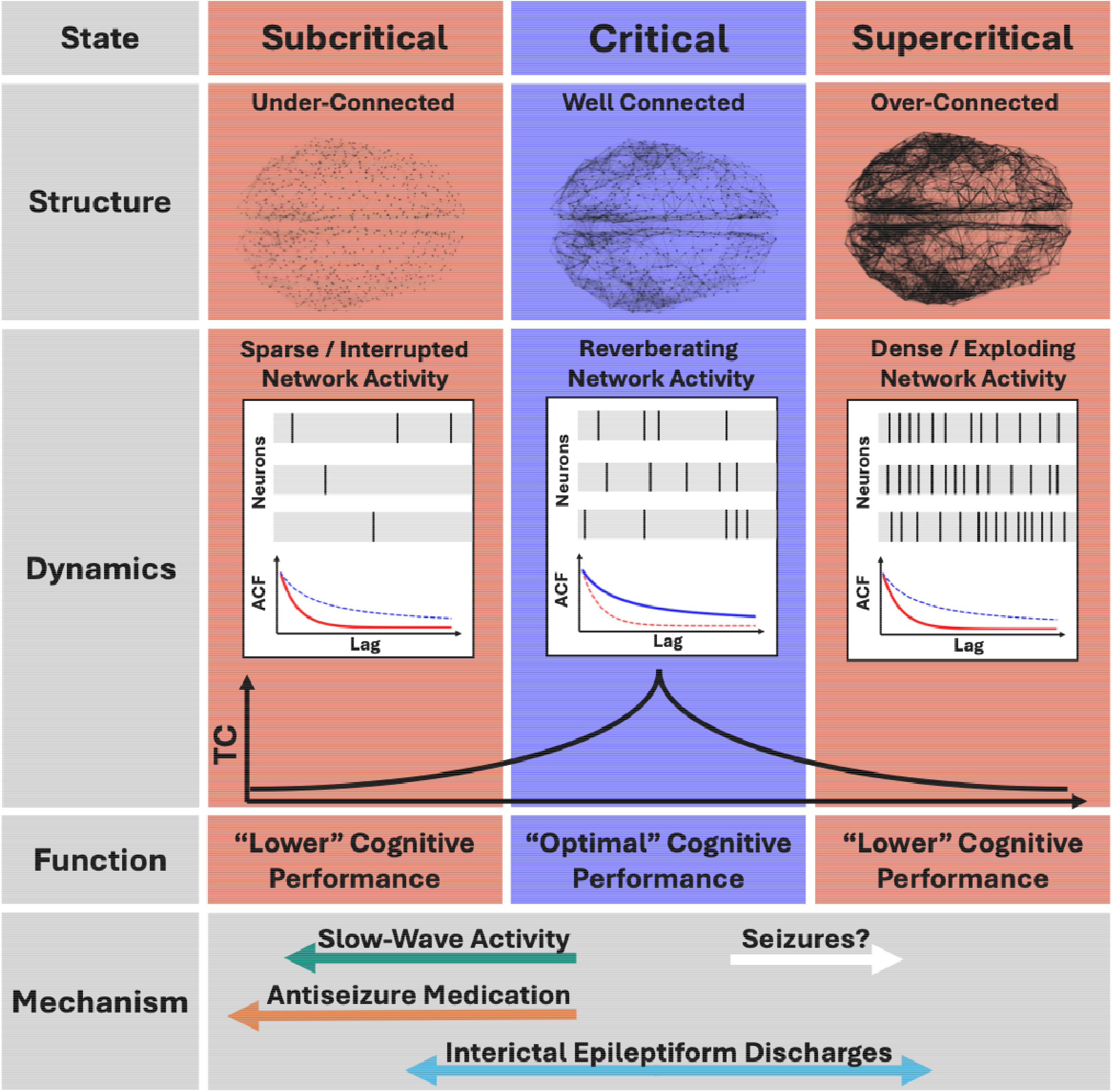
Illustration of the link between structure, dynamics and function in the brain criticality framework and the action of perturbative mechanisms. Subcritical: Under-connected networks showing sparse and interrupted activity do not allow for the propagation of information, resulting in short temporal correlations and lower cognitive performance. Critical: Well-connected networks with reverberating, critical dynamics exhibit maximized temporal correlations leading to optimal cognitive performance. Supercritical: Over-connected networks exhibiting dense activity lead to a fast decay of information through chaotic interactions. In this state temporal correlations are short and limited cognitive performance. We here presented evidence that low temporal correlations predict lower cognitive performance, and that three distinct mechanisms, slow-wave activity, antiseizure medication and interictal epileptiform discharges push the systems away from optimal critical dynamics.

**Table 1.**
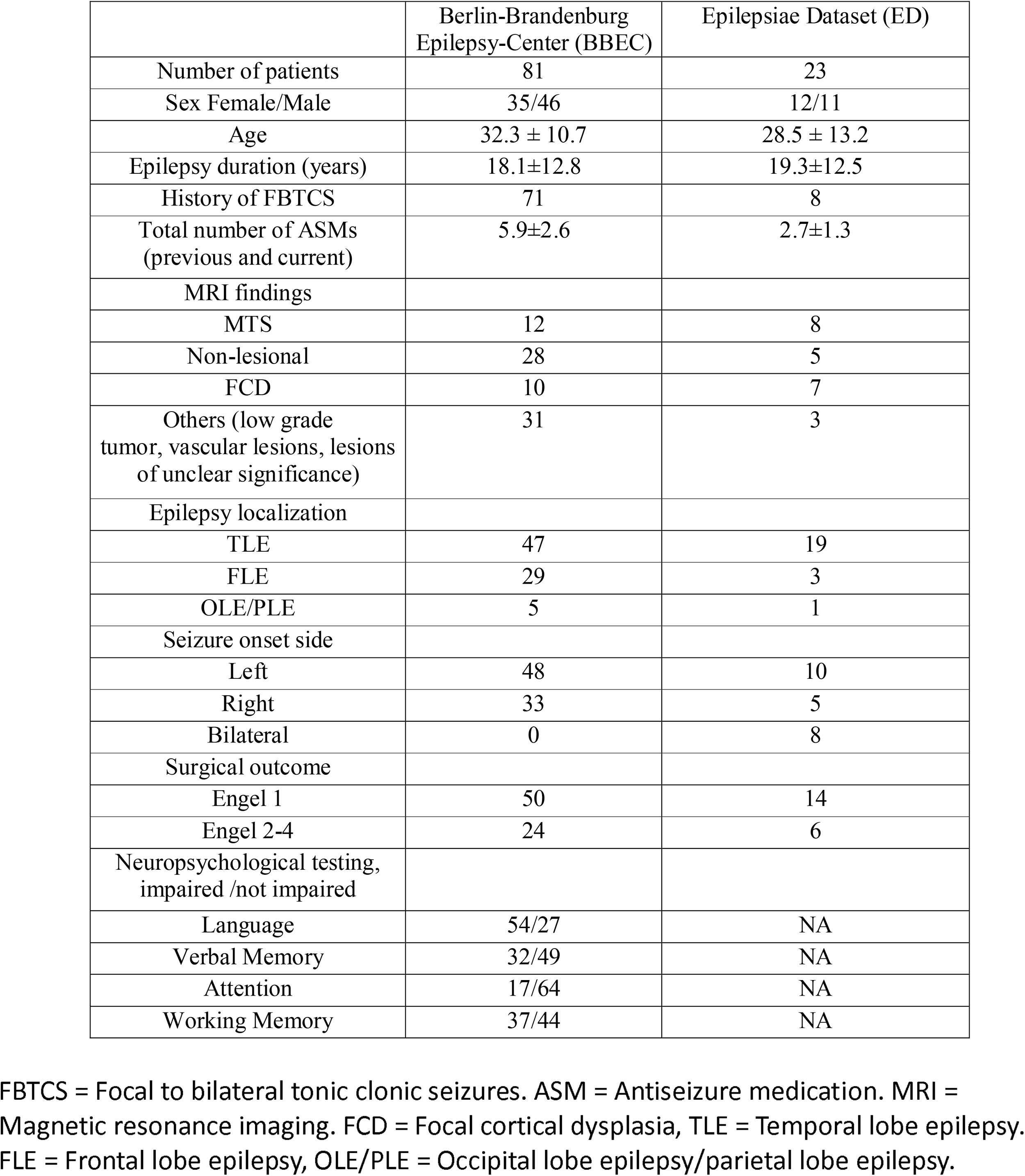
Patient characteristics.

|  | Berlin-Brandenburg<br>Epilepsy-Center (BBEC) | Epilepsiae Dataset (ED) |
| --- | --- | --- |
| Number of patients | 81 | 23 |
| Sex Female/Male | 35/46 | 12/11 |
| Age | 32.3 ± 10.7 | 28.5 ± 13.2 |
| Epilepsy duration (years) | 18.1±12.8 | 19.3±12.5 |
| History of FBTCs | 71 | 8 |
| Total number of ASMs<br>(previous and current) | 5.9±2.6 | 2.7±1.3 |
| MRI findings |  |  |
| MTS | 12 | 8 |
| Non-lesional | 28 | 5 |
| FCD | 10 | 7 |
| Others (low grade<br>tumor, vascular lesions, lesions<br>of unclear significance) | 31 | 3 |
| Epilepsy localization |  |  |
| TLE | 47 | 19 |
| FLE | 29 | 3 |
| OLE/PLE | 5 | 1 |
| Seizure onset side |  |  |
| Left | 48 | 10 |
| Right | 33 | 5 |
| Bilateral | 0 | 8 |
| Surgical outcome |  |  |
| Engel 1 | 50 | 14 |
| Engel 2-4 | 24 | 6 |
| Neuropsychological testing,<br>impaired /not impaired |  |  |
| Language | 54/27 | NA |
| Verbal Memory | 32/49 | NA |
| Attention | 17/64 | NA |
| Working Memory | 37/44 | NA |

FBTCs = Focal to bilateral tonic clonic seizures. ASM = Antiseizure medication. MRI = Magnetic resonance imaging. FCD = Focal cortical dysplasia, TLE = Temporal lobe epilepsy. FLE = Frontal lobe epilepsy, OLE/PLE = Occipital lobe epilepsy/parietal lobe epilepsy.

## Data Availability

Code for the model will be made available at https://gitlab.com/computational-neurologie/stc_model_2024 at the time of publication. Code for the analysis will be made available under https://gitlab.com/computational-neurologie/sde_n_cog for the BBEC dataset and https://gitlab.com/computational-neurologie/ieegCD for the ED. The ED dataset is publicly available within the Epilepsiae database.

https://gitlab.com/computational-neurologie/sde_n_cog

https://gitlab.com/computational-neurologie/ieegCD

http://www.epilepsiae.eu

