## Supplementary figures and images for "Critical dynamics predict cognitive performance and are disrupted by epileptic spikes, antiseizure medication and slow-wave activity"

### Supplementary Figure 1

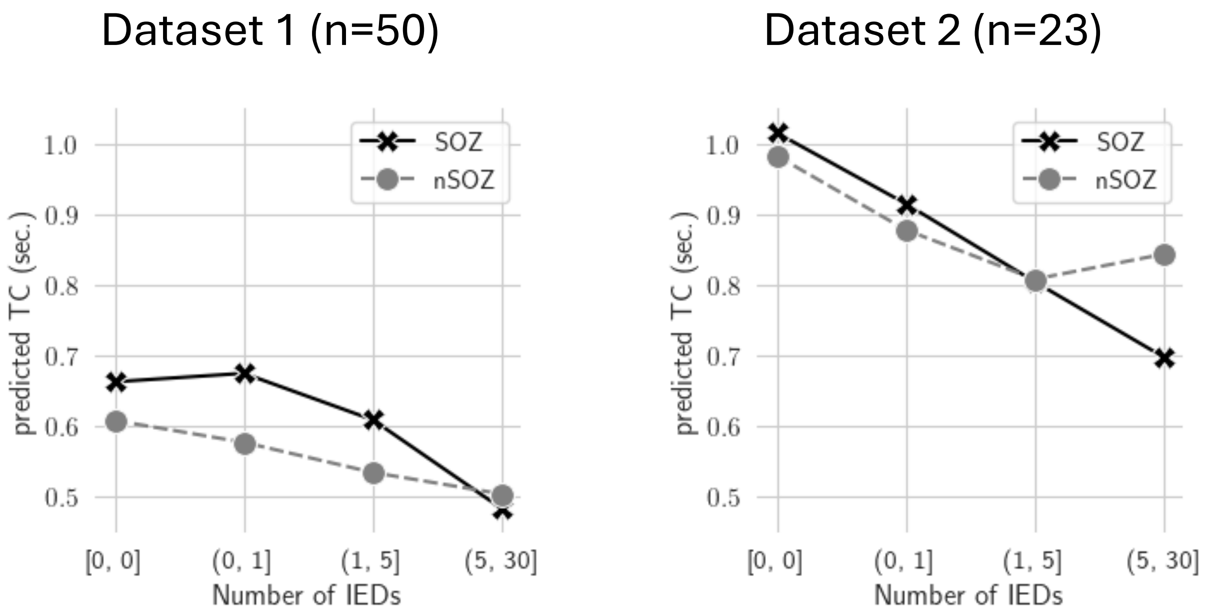
